# Radiomic signature accurately predicts the risk of metastatic dissemination in late-stage non-small cell lung cancer

**DOI:** 10.1101/2023.01.31.23285230

**Authors:** Agata Małgorzata Wilk, Emilia Kozłowska, Damian Borys, Andrea D’Amico, Krzysztof Fujarewicz, Izabela Gorczewska, Iwona Dębosz-Suwińska, Rafał Suwiński, Jarosław Śmieja, Andrzej Swierniak

## Abstract

**Background:** Non-small cell lung cancer (NSCLC) is the most common type of lung cancer, and the median overall survival is approximately 2-3 years among patients with stage III disease. Furthermore, it is one of the deadliest types of cancer globally due to non-specific symptoms and the lack of a biomarker for early detection. The most important decision that clinicians need to make after a lung cancer diagnosis is the selection of a treatment schedule. This decision is based on, among others factors, the risk of developing metastasis.

**Methods:** A cohort of 115 NSCLC patients treated using chemotherapy and radiotherapy with curative intent was retrospectively collated and included patients for whom positron emission tomography/computed tomography (PET/CT) images, acquired before radiotherapy, were available. The PET/CT images were used to compute radiomic features extracted from a region of interest, the primary tumor. Radiomic and clinical features were then classified to stratify the patients into short and long time to metastasis, and regression analysis was used to predict the risk of metastasis.

**Results:** Classification based on binarized metastasis-free survival (MFS) was applied with moderate success. Indeed, an accuracy of 0.73 was obtained for the selection of features based on the Wilcoxon test and logistic regression model. However, the Cox regression model for metastasis risk prediction performed very well, with a concordance index (c-index) score equal to 0.84.

**Conclusions:** It is possible to accurately predict the risk of metastasis in NSCLC patients based on radiomic features. The results demonstrate the potential use of features extracted from cancer imaging in predicting the risk of metastasis.

## Introduction

Lung cancer is one of the most frequently diagnosed cancer types worldwide, constituting over 11% of all cancer cases. With 2.2 million new diagnoses in 2020 alone, it was surpassed in incidence only by breast cancer, making the lung the most prevalent cancer site in men (with over 1.43 million diagnoses) and the third most prevalent in women after breast and colorectal cancers, which had 0.77 million diagnoses (1). While tobacco smoking is recognized as the primary cause of lung cancer, it can also be attributed to environmental factors such as air pollution, occupational exposure, and genetic predisposition (2–4). It is usually diagnosed at an advanced stage due to non-specific early-stage symptoms, which is reflected in the very high mortality rate. Indeed, the five-year survival rate for lung cancer does not exceed 20% (5–7), thus, it is the leading cause of cancer-related mortality and is responsible for 18% of all deaths from cancer (1).

Diagnosis of lung cancer involves medical imaging, including X-ray and positron emission tomography/computed tomography (PET/CT), which allows for classification according to the tumor node metastasis (TNM) staging system. Detected lesions are sampled by endobronchial ultrasound (EBUS) guided bronchoscopy and undergo histopathological assessment. Management is stage-specific (7), with clinical guidelines divided into early-stage, locally advanced, and metastatic cancer (8,9). In early-stage lung cancer, lobectomy is the preferred treatment option. If the tumor is not initially resectable, neoadjuvant chemotherapy can be implemented to downgrade the tumor, which would eventually allow for surgery. For selected patients with comorbidities, stereotactic radiotherapy (SABR) may also be considered. For locally advanced cancer with lymph node involvement, platinum-based chemotherapy administered concurrently or sequentially with radiotherapy is the most commonly used curative therapeutic option, and it can be followed by maintenance immunotherapy. For advanced metastatic cancer, immune checkpoint inhibitors, with or without chemotherapy, are a viable therapeutic option. As molecular diagnostics becomes routinely available, targeted therapies aimed at epidermal growth factor receptor (EGFR) (10), fibroblast growth factor receptor (FGFR) (11), anaplastic lymphoma kinase (ALK) (12), or Kirsten rat sarcoma virus (KRAS) (13) are being used to treat mutation carriers.

One of the main reasons for the high mortality seen in lung cancer is its invasiveness, and most patients develop distant metastases. Unfortunately, metastatic tumors are often resistant to treatment, which leads to much shorter survival times for these patients. Although the exact mechanisms of metastasis are still being investigated, it is known that cancer cells can spread by both blood and lymphatic vessels (14). Lung cancer metastases are most frequently observed in the brain, bones, liver, lung, and adrenal gland (15). Since the occurrence of distant metastasis is the turning point in the course of the disease, it might be considered an important endpoint in prognostic analysis, along with the standard endpoints. Furthermore, the ability to predict when lung cancer will metastasize could guide clinical decision-making and may be used to indicate the need for therapy intensification in high-risk patients.

The search for accurate prognostic biomarkers in lung cancer is hindered by its high heterogeneity and complexity. Nonetheless, clinical and molecular characteristics have shown some promise in predicting metastasis. Metastasis-associated lung adenocarcinoma transcript 1 (*MALAT-1*), a long non-coding ribonucleic acid (RNA), was demonstrated to be significantly associated with metastasis in non-small cell lung cancer (NSCLC) (16). Meanwhile, cancer antigen 125 (CA125) and neuron-specific enolase (NSE) were found to be indicative of liver metastasis (17). NSE, histological type, number of metastatic lymph nodes, and tumor grade were used to construct a nomogram for use in brain metastasis prediction (18). Vimentin expression was also identified as a potential predictor of brain metastasis in *EGFR*-mutant NSCLC patients (19).

Recently, medical imaging has gained attention as an alternative source of biomarkers (20–22). It has distinct advantages over molecular markers in that it is non-invasive, requires no additional assays, and utilizes information acquired during a routine diagnostic procedure. As such, imaging biomarkers are also the fastest to obtain, making them perfect for therapy planning. Two main strategies can be used to acquire biomarkers. The first is to directly analyze raw images. Another solution is radiomics, in which segmented images are subjected to feature extraction. This method provides numerical variables that describe the shape and texture of the region of interest (ROI), which can then be used in statistical or machine-learning models.

The radiomics-based approach has been successfully applied for different endpoints in lung cancer, including overall survival (OS) and progression-free survival (PFS) (23). It has also shown promising results for the prediction of distant metastases. Coroller et al. (24) selected a radiomic signature based on CT images to predict distant metastasis in lung adenocarcinoma. Wu et al. constructed and validated a Cox proportional hazards model using 18F-fluorodeoxyglucose PET (^18^F-FDG PET) imaging to predict freedom of distant metastasis in early-stage NSCLC patients (25). Fave et al. (26) demonstrated that adding pre-treatment radiomic features extracted from CT images could improve the ability of clinical prognostic models to predict distant metastasis (26). Meanwhile, Dou et al. (27) focused on locally advanced lung adenocarcinoma and investigated radiomic features from the primary tumor and peritumoral region (27).

In this study, 115 NSCLC patients with various histological subtypes were retrospectively analyzed. The prognostic value of standard clinical features, and radiomic features extracted from PET/CT images acquired for radiotherapy planning, were evaluated by determining if they could be used to predict time to distant metastasis. To answer this question, machine learning models were constructed for continuous and categorical metastasis-free survival (MFS) prediction.

## Materials and Methods

### Study design

A cohort of NSCLC patients was collated to investigate if PET/CT imaging routinely performed for radiotherapy planning could help in planning the future treatment strategy, with a focus on predicting the risk and time of relapse with distant metastases. MFS was defined as the time elapsed between diagnosis and the detection of distant metastasis or the time of death/last follow-up if distant metastases did not emerge. In addition, classification algorithms were used to predict if MFS would be short or long.

As the prediction of metastasis risk was the focus of the study, the primary lung cancer tumor was the ROI. Using the available PET/CT scans, radiomic features were extracted from the ROI and assessed.

The specific clinical question considered in this work was whether or not a radiomic signature could be extracted that would help discriminate between a primary tumor that has the potential to metastasize early from one that metastasizes late or not at all.

### Study population

Data were collected retrospectively at the Maria Sklodowska-Curie National Research Institute of Oncology, Gliwice Branch (NRIO). The cohort consisted of 115 patients with NSCLC who were treated with curative intent at the Institute between 2009 and 2017. All patients in the cohort had been treated with a combination of chemotherapy and radiotherapy. Most of the patients received a platinum-based doublet with vinorelbine. Patients received between one and six cycles (median four), followed by radiotherapy (RT) with a total dose between 60 and 70 Gray (Gy) in two Gy fractions. The study was approved by the Local Bioethical Committee of the NRIO in accordance with national regulations. Formal written consent was obtained from all participants of the study. The clinical data were anonymized before the computational analysis.

All patients underwent PET/CT imaging for radiotherapy planning. Only patients with nondetectable distant tumors at the onset of treatment were assessed. However, most patients had locally disseminated tumors to the lymph nodes, as they were diagnosed late due to non-specific symptoms.

In the cohort, 72% of patients were males, and 28% were female. This is consistent with population data showing that most lung cancer patients are male. The median age of patients in the cohort was 61 years, and over half of the patients had tumors located in the left lung. The most prevalent cancer subtype was squamous cell carcinoma, which constituted two-thirds of all cases, followed by large cell carcinoma (24.3%) and adenocarcinoma (7.0%). Detailed characteristics of the cohort are presented in Table 1.

**Table 1.** Patient characteristics. For continuous variables, median and quartiles are listed.

|  |  | N = 115 |
| --- | --- | --- |
| Sex | Male | 83 (72.2%) |
|  | Female | 32 (27.8%) |
| **Age** |  | 61 (57-67) |
| **Histopathology** | Squamous | 77 (67.0%) |
|  | Adenocarcinoma | 8 (7.0%) |
|  | Large cell | 28 (24.3%) |
|  | Other | 2 (1.7%) |
| **Location** | Left | 65 (56.5%) |
|  | Right | 50 (43.5%) |
| **T** | 1 | 4 (3.5%) |
|  | 2 | 37 (32.2%) |
|  | 3 | 37 (32.2%) |
|  | 4 | 37 (32.2%) |
| **N** | 0 | 19 (16.5%) |
|  | 1 | 6 (5.2%) |
|  | 2 | 83 (72.2%) |
|  | 3 | 7 (6.1%) |
| **M** | 0 | 115 (100%) |
|  | 1 | 0 (0%) |
| **Zubrod score** | 0 | 34 (29.6%) |
|  | 1 | 80 (69.6%) |
|  | 2 | 1 (0.9%) |

The median time-to-metastasis was 2.77 years, with a secondary tumor observed most frequently in the second lung, brain, bones, and liver.

### Positron emission tomography/computed tomography data acquisition and segmentation

The PET/CT images were acquired at the NRIO using Philips GeminiGXL 16 (Philips, Amsterdam, Netherlands) (24 patients) and Siemens Biograph mCT 131 (Siemens AG, Munich, Germany) (88 patients) PET/CT scanners. For each patient, the ROI was contoured by the same experienced nuclear medicine specialist using Medical Image Merge (MIM) 7.0.1 software and the PET Edge™ tool (both MIM Software Inc., OH, USA).

### Extraction of radiomic features

Feature extraction was performed with PyRadiomics version 3.0.1, a Python package designed to increase the reproducibility of radiomic studies (28). Using the PET dataset, 105 standard features were calculated. Radiomic features belong to one of three classes, including first-order statistics such as energy, entropy, and minimum, as well as shape features such as volume, surface area, and sphericity, and texture features including Gray Level Co-occurrence Matrix (GLCM), Gray Level Dependence Matrix (GLDM), Gray Level Run Length Matrix (GLRLM), Gray Level Size Zone Matrix (GLSZM), and Neighboring-Gray Tone Difference Matrix (NGTDM).

### Metastasis-free survival categorization

For the classification, a threshold of one year was used to create two classes, which included patients with MFS below and over this threshold. Due to the presence of censored observations (in the cohort this primarily signified the patient’s death), such stratification divided patients into a group who suffered either metastasis or death within a year (66 patients), and those who did not (49 patients). To create subgroups that were more related to the research question, the binary MFS was defined as “short” if the patient developed metastasis within a year (25 patients) and “long” if the patient developed metastasis or was censored after longer than a year (49 patients).

### Statistical analysis

Statistical analysis was performed using the R environment (version 4.1.3). For survival analysis, survival (version 3.2-13) was used, caret (version 6.0.93) and RandomForest (version 4.7-1.1) were used for classification, and randomForestSRC (version 3.1.1) was used to perform random survival forest. A heatmap of the radiomic features was created with ComplexHeatmap (version 2.10.0).

Filtering of the radiomic features was applied based on the Pearson correlation coefficient to avoid redundancy, with a cutoff threshold equal to 0.9 (see Supplementary Table 1). Since the PET images were acquired using two scanners, principal component analysis was applied to determine if there was any grouping of samples due to the scanner used (see Supplementary Figure 1).

The clinical and radiomic features with potential for event-free survival (EFS) and MFS prediction were assessed (see Supplementary Table 2). In addition, differences in the values of radiomic and clinical features between ‘short’ and ‘long’ MFS patient subgroups were investigated statistically. Fisher’s exact test was performed for categorical variables, while the MannWhitney U test was used for continuous variables (see Supplementary Table 3). A log-rank test was also conducted for both categorical and continuous features (see Supplementary Table 4). As the log-rank test assesses if there is a significant difference between two or more survival curves, continuous features were binarized with respect to the median value.

### Cross-validation

The value of any type of predictive model lies in its applicability to unknown data, and not just its ability to fit the training data. Cross-validation enables evaluation of the model’s ability to generalize by removing part of the data from the cohort and applying them in the estimation of model performance. In addition, data partitioning at the beginning of each iteration prevents information leakage.

For a more consistent comparison between the regression and classification results, modified k-fold data partitioning was applied. Firstly, the data was ordered according to (continuous) MFS values. Then, the observations were assigned consecutive numbers, from one to five, which were used as cross-validation folds. Such partitioning ensures proper stratification of both continuous and binarized MFS.

### Classification algorithms

The observed relationships between binary MFS and binary EFS and extracted features (both clinical radiomic) were verified by employing classification models. Firstly, three main feature selection methods were applied, including Student’s t-test, Wilcoxon test, and a mutual information test. To investigate the impact of a varying number of features on classification quality, between 1 and 10 features were tested. Since only the mutual information method handles both categorical and continuous variables, a hybrid selection was used for the other two methods by applying the main method for continuous variables and Fisher’s exact test for categorical variables. The categorical variables that passed the significance threshold equal to 0.1 were added to the model.

The following classification methods were tested: K Nearest Neighbor (KNN) with different K values (for clarity, only the best one, K=5, is presented), random forest, support vector machines (SVM) with linear and radial kernels, and logistic regression (LogReg). Considering the inconsistent orders of magnitude for radiomic features, a z-score transformation was used to scale the data. In each k-fold iteration, the scaling parameters (mean and standard deviation) were determined from the training set and applied to both the training and test sets. Classification accuracy was then used to assess model performance.

### Regression algorithms

For the prediction of continuous MFS, Cox proportional hazards regression (using survival R package) and random survival forest (using randomForestSRC R package) were applied. Variable selection was performed based on univariate analysis, with the Harrell Concordance index (C-index) adopted as a ranking metric. The model performance was validated using the k-fold partitioning described above. Again, models containing between 1 and 10 features were tested.

### Radiomic-based risk score

Although cross-validation facilitates the estimation of prediction quality, the results and selected features can be different in each iteration due to subsampling. Therefore, all selections were repeated on the entire dataset to obtain conclusive feature rankings. To demonstrate the validity of the obtained signature, the Cox model was chosen, which is the classic approach to survival data analysis with known interpretation. The patients were then divided into high-risk and low-risk groups based on the calculated median risk score, and MFS was compared using Kaplan-Meier curves.

## Results

### Patient characteristics

The cohort included only NSCLC patients, as it is the most common type of lung cancer. Most patients (67%) had squamous histopathological subtypes, and almost two-thirds had an advanced stage of the primary tumor (T3 or T4). In total, 37 patients eventually developed distant metastases. Figure 2A shows a Kaplan-Meier plot for MFS probability in the entire patient cohort.

**Figure 1:**
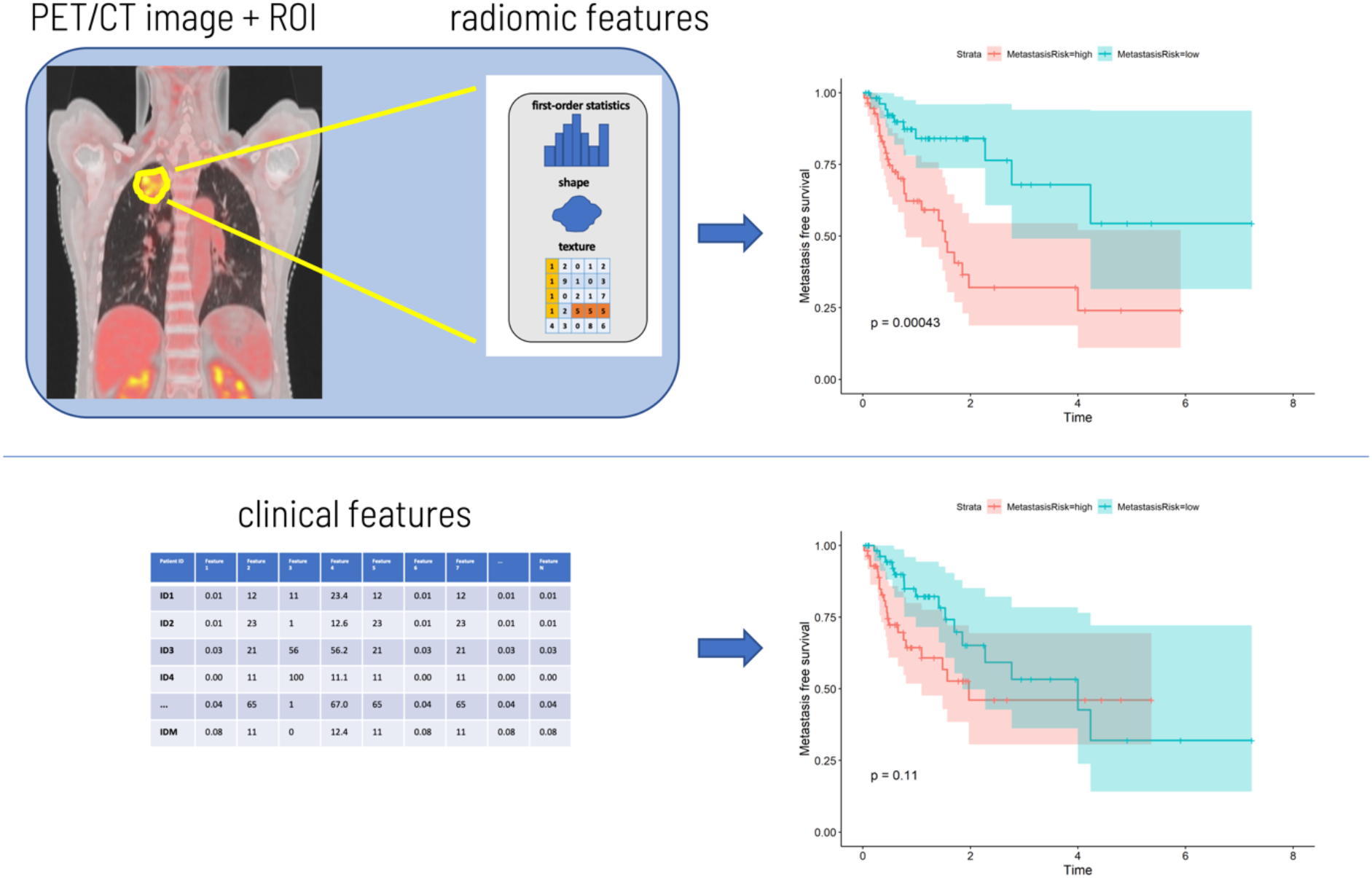
Project workflow. Positron emission tomography/computed tomography (PET/CT) images were acquired and radiomic features were extracted from regions of interest (ROI). Integration of clinical and radiomic data led to the prediction of short-term and long-term metastasis-free survival (MFS) and the risk of metastasis. The output from the workflow was a radiomic signature, which could be used for the prediction of metastasis risk in newly diagnosed NSCLC patients being treated with platinum-based chemotherapy.

**Figure 2:**
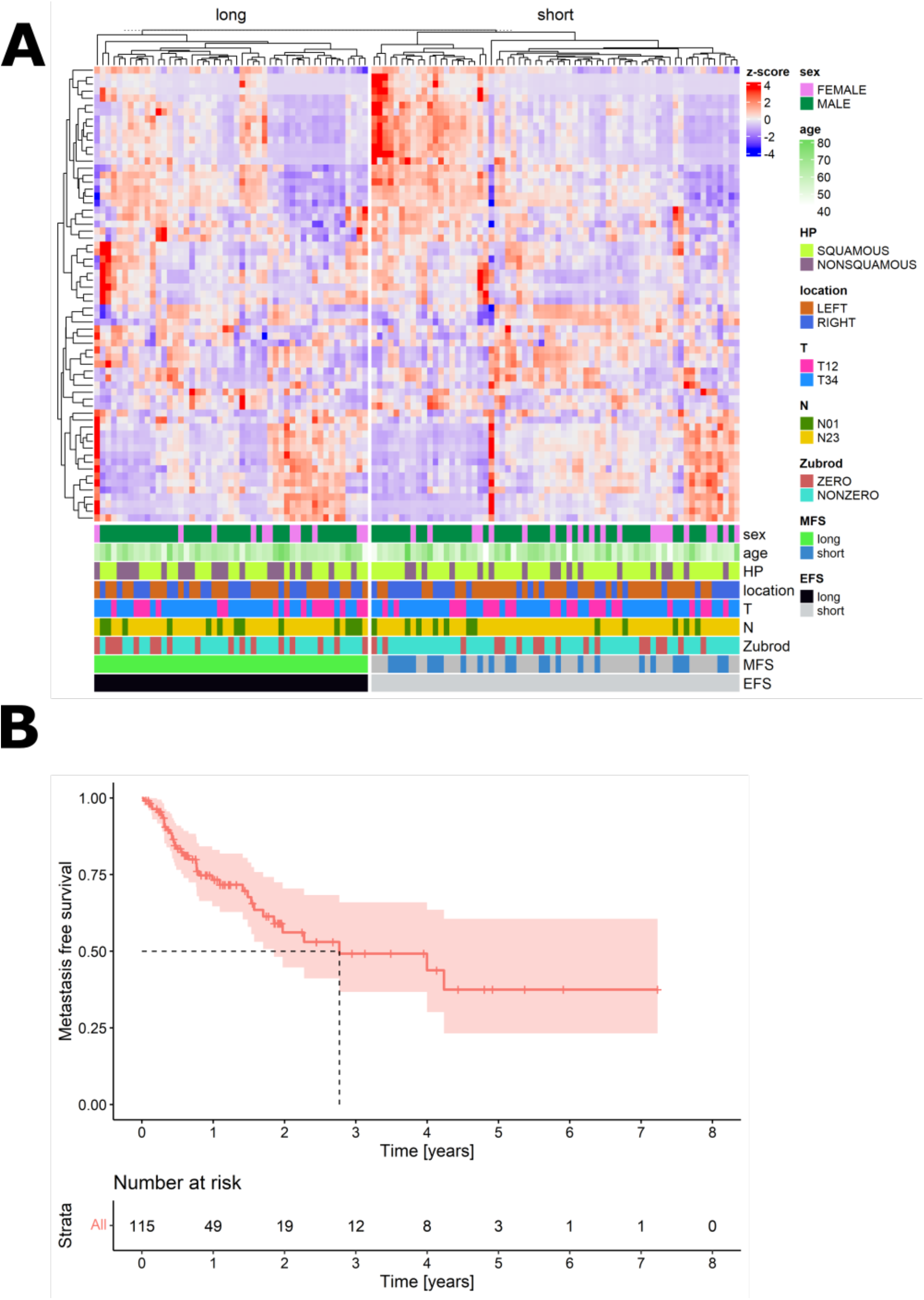
The integration of clinical and radiomic data. A. Kaplan-Meier plot of metastasisfree survival (MFS) for the entire population. B. Integration of clinical and radiomic data. Patients were split by the binary event-free survival (EFS).

**Figure 3:**
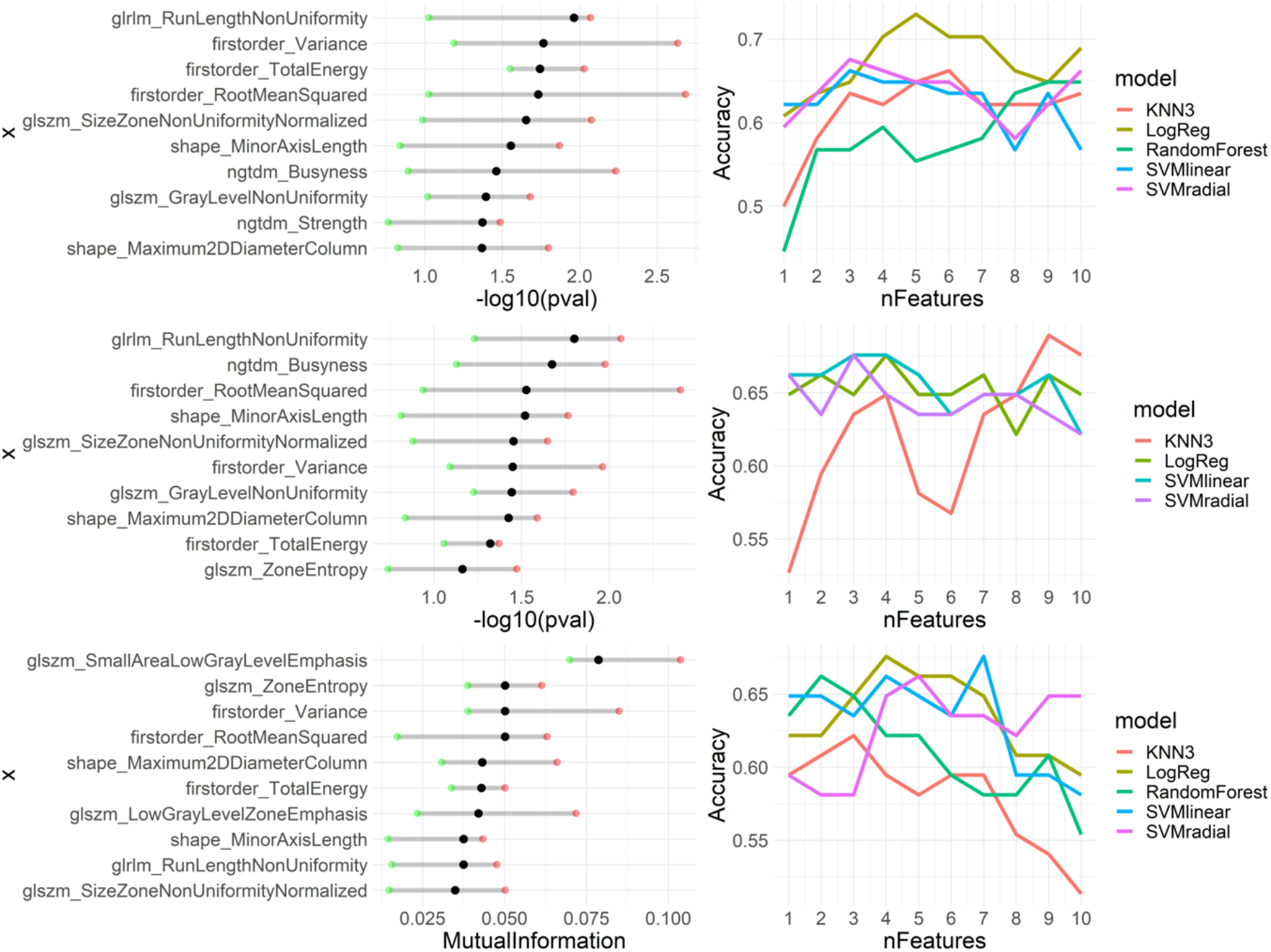
Metastasis-free survival (MFS) prediction using the classification approach. Top row: Wilcoxon test; middle row: Student’s t-test; bottom row: mutual information test. Left column: feature selection in a 5-fold cross-validation. Features were ranked according to the log10(p-value) for the Wilcoxon test and Student’s t-test selections, and mutual information score for mutual information selection. Black dots indicate the median value across folds, green dots indicate the lowest value across folds, and red dots indicate the highest value. Right column: classification results for the test set in a 5-fold cross-validation for different models, depending on the number of features.

**Figure 4:**
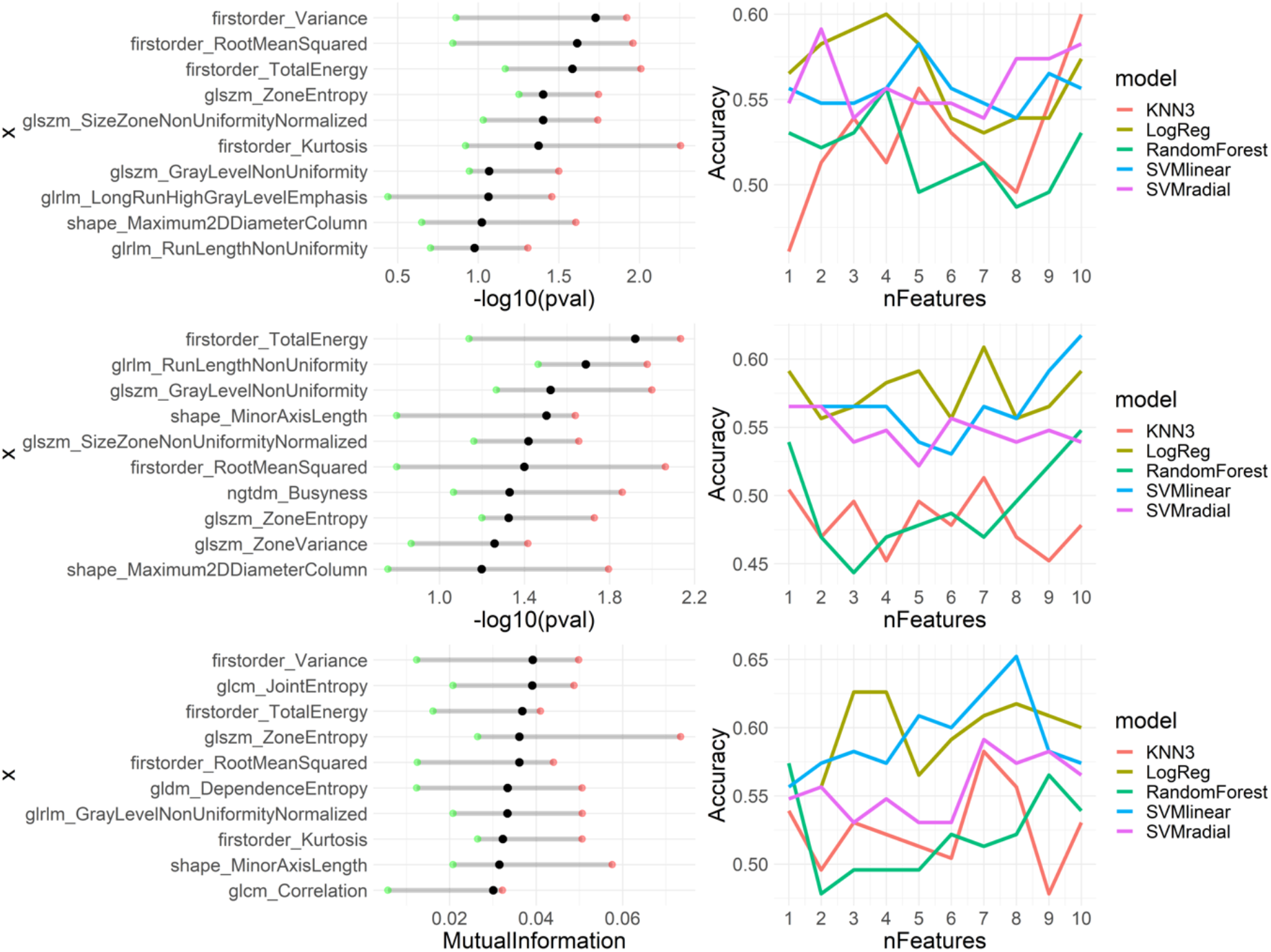
Event-free survival (EFS) prediction using the classification approach. Top row: Wilcoxon test; middle row: Student’s t-test; bottom row: mutual information. Left column: feature selection in a 5-fold cross-validation. Features are ranked according to the -log10(p-value) for the Wilcoxon test and Student’s t-test selections, and mutual information score for mutual information selection. Black dots indicate the median value across folds, green dots indicate the lowest value across folds, and red dots indicate the highest value across folds. Right column: classification results for the test set in a 5-fold cross-validation for different models, depending on the number of features.

**Figure 5:**
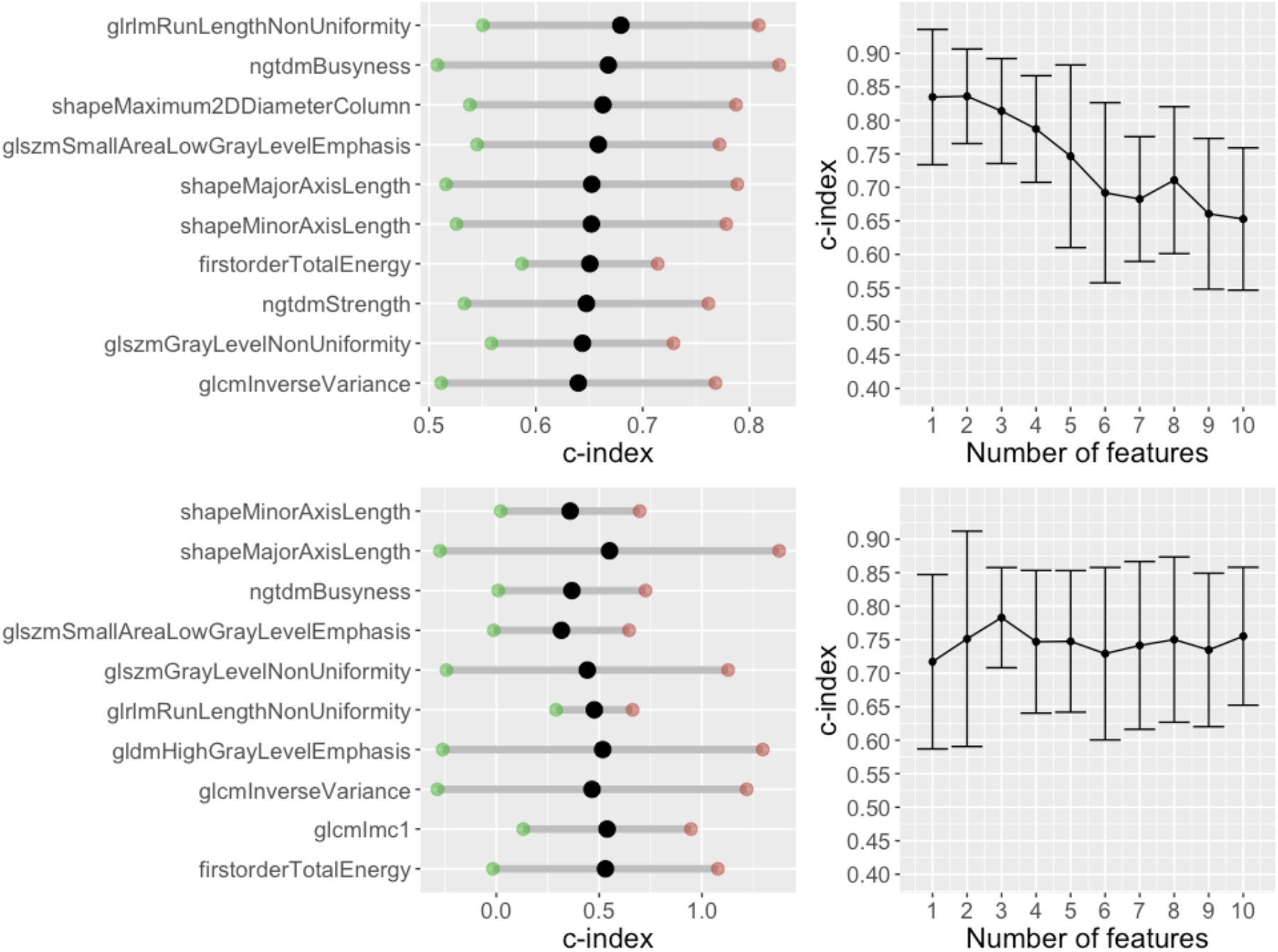
Metastasis-free survival (MFS) prediction using a regression approach. Top row: Cox regression, bottom row: random survival forest. Left column: feature selection in a 5-fold crossvalidation. Features were ranked according to the concordance index value for the univariate model. Right column: Prediction results for the test set in a 5-fold cross-validation, depending on the number of features.

None of the clinical features of the cohort were informative in relation to the time to metastasis onset (Supplementary Table 2). This means that clinicians are unable to predict if a particular patient will develop metastatic cancer, based only on clinical variables at diagnosis. On the other hand, 34 radiomic features were statistically significant against continuous MFS, 36 features against the binarized MFS, and 18 against EFS.

### Integration of clinical and radiomic data

High correlations were observed between the radiomic features, which resulted in only 65 of 105 features passing the initial correlation filtering. The highest redundancy was found for the first-order features (6 out of were18 kept) and the lowest for the GLSZM features (15 out of 16 were kept) (see Supplementary Table 1).

Correlations between radiomic and clinical features were mostly low, signifying that both datasets carried independent information. Also, the hierarchical clustering of radiomic features did not correspond to any discernible grouping of clinical features (see Figure 2).

Figure 2B shows the normalized z-score values of radiomic features for each patient. The patients were divided into short and long EFS groups. As can be seen from the results, the hierarchical clustering correctly divided patients into these two groups. Furthermore, it was observed that the radiomic feature spectrum varied between patients with short and long EFS. This demonstrates that there is potential for the use of radiomic features in predicting EFS.

### Classification of advanced non-small cell lung cancer

As expected, no clinical features were selected by the models. The feature rankings obtained for EFS and MFS prediction differed, which aligns with the different interpretations of these endpoints. While the rankings varied with respect to feature selection and classification methods, there was some consistency among the top features. Indeed, TotalEnergy, ZoneEntropy, and RootMeanSquared favored EFS prediction, while Variance, TotalEnergy, RunLengthNonUniformity (GLRLM), SizeZoneNonUniformityNormalized, and Maximum2DDiameterColumn favored MFS prediction.

The highest accuracy for EFS prediction (approx. 0.65) was achieved using the SVM classifier with linear kernels for the mutual information selection of eight features. The highest accuracy for MFS prediction (approx. 0.73) was achieved using the LogReg classifier for the five features selected using the Wilcoxon test. Due to the imbalanced classes, with “long” (treated as the negative class) being the predominant group, the models for MFS tended to yield high specificity and relatively low sensitivity. Most models performed better for a small number of features.

### Prediction of risk of metastasis

For regression-based models, the tendency was similar, with the highest predictive ability observed for a small number of features. The highest median C-index across folds was reached for two features (GLRLM and NGTDM Business) in Cox regression and one feature (shapeMinorAxisLength) in the random survival forest.

The mean C-index for the best set of features using Cox regression was 0.84, whereas the Cindex for the random survival forest was 0.8. The inclusion of more features in the model resulted in a loss of prediction quality due to overfitting. No clinical features were selected for the best models, which is consistent with the preliminary patient cohort analysis.

Feature selection on the entire dataset revealed that the two top features for Cox regression, SmallAreaLowGrayLevelEmphasis (GLSZM) and GLRLM, also held high-ranking positions in the classification approach. Therefore, the Cox model was constructed using those two features. The high-risk and low-risk groups (Figure 6) had significantly different MFS, with the log-rank test *P*<0.001.

**Figure 6:**
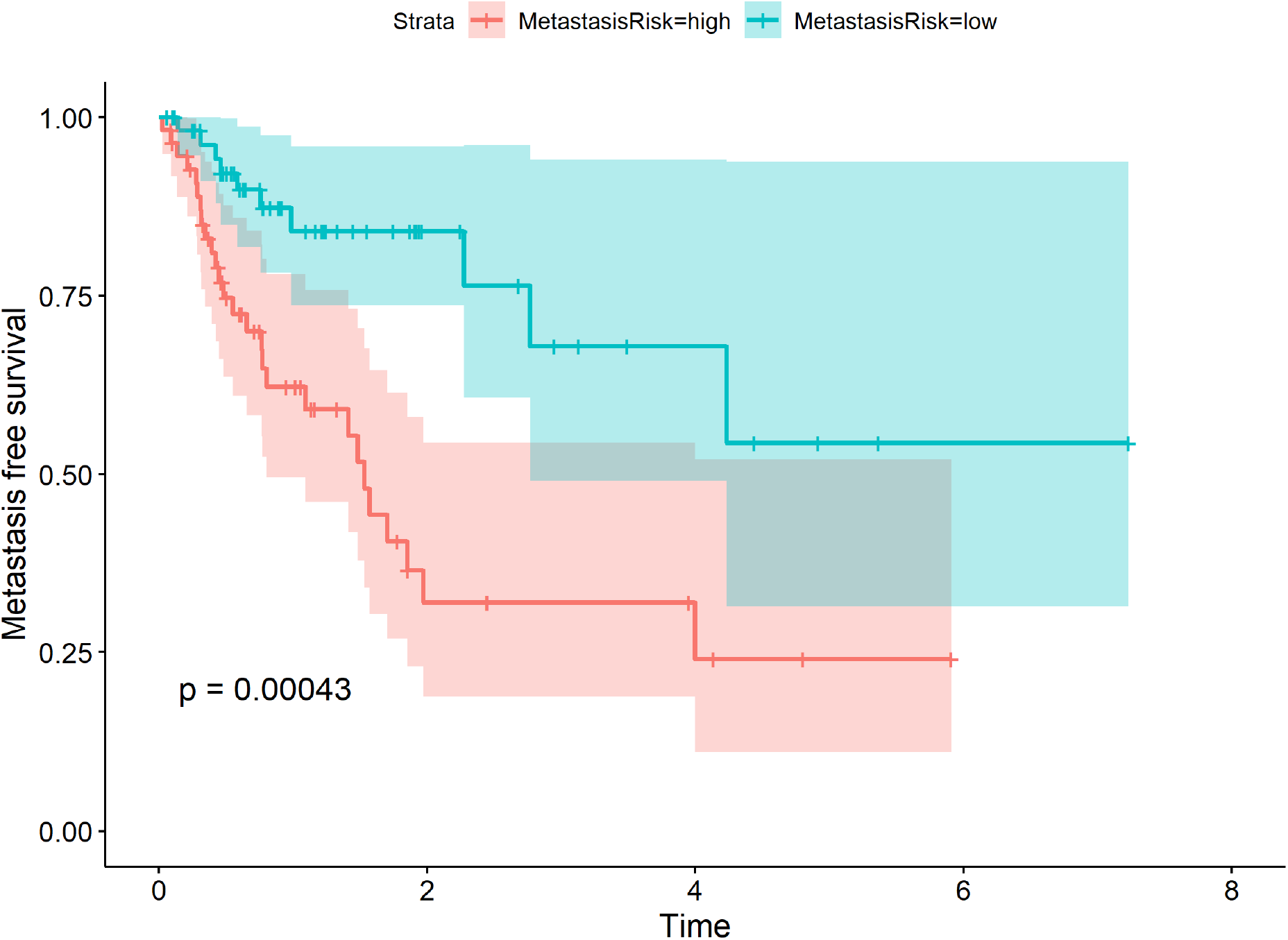
Kaplan-Meier plot of metastasis-free survival for the whole cohort. The patients were divided into high-risk and low-risk groups according to a Cox model constructed using the feature selection and number for which the cross-validation accuracy was highest.

## Discussion

Lung cancer is the leading cause of cancer-related death worldwide, claiming over 1.7 million lives yearly. It is characterized by high invasiveness, and the occurrence of distant spread significantly influences survival and treatment options. This necessitates the search for prognostic biomarkers that could help determine the time to metastasis onset. With the rapid development of the radiomics field, researchers have turned to medical imaging, which is routinely performed and non-invasive, as a source of information that could shed some light on the tumor dissemination process and aid clinicians in therapy planning.

A cohort of NSCLC patients with different subtypes and stages of the disease was collated. It was concluded that the standard clinical data available for the patients, except for higher metastatic potential exhibited by the squamous subtype, were largely uninformative regarding metastasis occurrence. To assess the potential of radiomics for MFS prediction, we extracted 105 radiomic features from PET/CT scans, using the primary tumor as the ROI. Regression and machine learning methods were then used to select radiomic signatures that could predict the risk of metastasis and achieved a C-index of 0.84 for the Cox proportional hazards model and 0.8 for the random survival forest, and an accuracy of 0.72 for the KNN classifier. These results confirm that medical images contain information that could be successfully applied to MFS prediction.

Several studies have shown the potential of radiomic features in predicting distant metastasis in lung cancer, with most of them focusing on either a particular subtype or stage. Coroller et al. (24) investigated radiomic features extracted from CT images for predicting distant metastasis in lung adenocarcinoma, which had a C-index of 0.61 on an independent validation set. Fave et al. (26) demonstrated that combining pre-treatment radiomic features with clinical information improved the ability of prognostic models to predict distant metastasis in stage III NSCLC patients, reporting a C-index of 0.63 (24). Wu et al. used features extracted from PET images to predict the freedom of distant metastasis, with a high C-index of 0.71 in independent validation (25). However, this work only focused on early-stage lung cancer. Dou et al. (27) presented an interesting approach, extracting features from both the tumor and tumor rim and achieving a C-index of 0.64 in a cohort of patients with locally advanced lung adenocarcinoma (27). In the current work, significantly better model quality was achieved in a cohort including patients with varying subtypes (squamous cell carcinoma, adenocarcinoma, large cell carcinoma) and stages.

While a regression approach, such as a Cox proportional hazards model, is typically used for survival-type analysis, the risk score it yields does not directly translate to the time of event occurrence. The C-index only compares pairs of observations, resulting in a global assessment of whether a higher risk is related to a shorter time-to-event. Therefore, classification was also performed and achieved an accuracy of 0.72 in cross-validation.

After testing several methods and approaches to variable selection, it was observed that similar predictive ability could be achieved for different feature sets, which indicates that even unrelated radiomic features carry equivalent information. Interestingly, the quality dropped drastically with increased feature numbers in all models. This suggests that features with high predictive potential perform much worse when combined than when used in isolation, and emphasizes the importance of selecting algorithms that are sensitive to feature interactions.

Certain variables retained high positions across different selections. These included GLRLM, NGTDM Strength, and NGTDM Business. This demonstrates that these radiomic features are important for predicting if and when metastasis will occur in a lung cancer patient.

This analysis was not without limitations. While the study design ensured all images were contoured by one expert, which prevented bias, this did not allow for an assessment of the reproducibility of radiomic feature extraction. In addition, plans are in place to collect an independent patient cohort to validate the signature. Future work will also investigate tumor growth and dissemination dynamics, to achieve more clinically meaningful predictions.

## Conclusions

Based on a cohort comprised of 115 NSCLC patients, clinical features routinely collected during diagnostic procedures are not sufficient for the prediction of the risk of metastasis. Medical images (PET/CT scans) were investigated as a potential source of prognostic markers by assessing radiomic features in various classes of predictive models. A model based on two texture features (GLSZM and GLRLM) was constructed, which divided the patient cohort into lowrisk and high-risk groups that significantly differed in MFS. The findings of this study have the potential to help clinicians make adjustments to therapy and create a rational basis for the intensification of systemic treatment in high-risk lung cancer patients.

## Data Availability

All data produced in the present study are available upon reasonable request to the authors

## Acknowledgments

This work was supported by the Polish National Science Centre, Grant Number: UMO-2020/37/B/ST6/01959, and Silesian University of Technology statutory research funds. Calculations were performed on the Ziemowit computer cluster in the Laboratory of Bioinformatics and Computational Biology, created in the EU Innovative Economy Programme POIG.02.01.00-00-166/08 and expanded in the POIG.02.03.01-00-040/13 project. Data analysis was partially carried out using the Biotest Platform developed within project PBS3/B3/32/2015, which was financed by the Polish National Centre of Research and Development (NCBiR). This work was carried out in part by the Silesian University of Technology internal research funding (A.M.W., E.K., D.B., K.F., J.S., and A.S.). The founders have no role in designing the study and writing the manuscript.

## Footnotes

### Reporting checklist

The authors have completed the “**Prediction Model Development and Validation**” reporting checklist.

### Data Sharing Statement

The authors submitted a data-sharing statement along with the manuscript.

### Ethical statement

The authors are accountable for all aspects of the work and will ensure that questions related to the accuracy or integrity of any part of the work are appropriately investigated and resolved. The study was conducted in accordance with the Declaration of Helsinki (as revised in 2013). The study was approved by the institutional board of Maria Skłodowska-Curie National Research Institute of Oncology (Gliwice Branch), and individual consent for this retrospective analysis was waived.

## Supplementary appendix

**Supplementary Table 1**. Summary of the correlation-based feature filtering.

**Supplementary Table 2**. Potential of the data to predict EFS (clinical variables). Fisher’s exact test was applied for p-value estimation.

**Supplementary Table 3**. Potential of the data to predict MFS and EFS. For binary MFS and EFS, the Mann-Whitney U test was used. For the continuous MFS log-rank test. Variables statistically significant against continuous MFS are highlighted in bold.

**Supplementary Table 4**. Log-rank test for continuous MFS.

**Supplementary Figure 1**. Principal component analysis of the radiomic features (after correlation filtering). Colors correspond to the PET/CT scanner. There is no visible grouping of samples according to the scanner.

**Supplementary Figure 2**. Correlation between radiomic features.

**Supplementary Figure 3**. Kaplan-Meier plot for metastatic-free survival with high/low SmallAreaLowGrayLevelEmphasis value

**Supplementary Figure 4**. Kaplan-Meier plot for metastatic-free survival with high/low Run-LengthNonUniformity value

