## Supplementary Appendix for "Radiomic signature accurately predicts the risk of metastatic dissemination in late-stage non-small cell lung cancer"

#### Supplementary Tables

**Supplementary Table 1.** Summary of the correlation-based feature filtering.

| Feature class | All features | Features kept after correlation filtering |
|---|---|---|
| First order | 18 | 6 |
| GLCM | 22 | 13 |
| GLDM | 14 | 8 |
| GLRLM | 16 | 10 |
| GLSZM | 16 | 15 |
| NGTDM | 5 | 4 |
| Shape | 14 | 9 |
| Total | 105 | 65 |

**Supplementary Table 2.** Potential of the data to predict EFS (clinical variables). Fisher's exact test was applied for p-value estimation.

| | | EFS short (N = 66) | EFS long (N = 49) | MFS short (N = 25) | MFS long (N = 49) | p-value (binary EFS) | p-value (binary MFS) |
|---|---|---|---|---|---|---|---|
| Sex | MALE | 45 | 38 | 17 | 38 | 0.299 | 0.408 |
| | FEMALE | 21 | 11 | 8 | 11 | | |
| HP | SQUAMOUS | 49 | 28 | 16 | 28 | 0.071 | 0.624 |
| | NONSQUAMOUS | 17 | 21 | 9 | 21 | | |
| Location | LEFT | 38 | 27 | 14 | 27 | 0.850 | 1 |
| | RIGHT | 28 | 22 | 11 | 22 | | |
| T | T12 | 24 | 17 | 8 | 17 | 1 | 1 |
| | T34 | 42 | 32 | 17 | 32 | | |
| N | N01 | 12 | 13 | 5 | 13 | 0.361 | 0.582 |
| | N23 | 54 | 36 | 20 | 36 | | |
| Zubrod score | ZERO | 17 | 17 | 6 | 17 | 0.311 | 0.431 |
| | NONZERO | 49 | 32 | 19 | 32 | | |

**Supplementary Table 3.** Potential of the data to predict MFS and EFS. For binary MFS and EFS, the Mann-Whitney U test was used. For the continuous MFS log-rank test. Variables statistically significant against continuous MFS are highlighted in bold.

| Feature | p-value (MFS) | p-value (EFS) | p-value (continuous MFS) |
|---|---|---|---|
| **original_glcm_InverseVariance** | **0.066** | **0.401** | **<0.001** |
| **original_glszm_SizeZoneNonUniformityNormalized** | **0.016** | **0.022** | **0.003** |
| **original_glszm_SmallAreaLowGrayLevelEmphasis** | **0.025** | **0.593** | **0.003** |
| **original_glrlm_LongRunLowGrayLevelEmphasis** | **0.071** | **0.555** | **0.004** |
| **original_shape_SurfaceVolumeRatio** | **0.061** | **0.115** | **0.005** |
| **original_glszm_GrayLevelNonUniformity** | **0.023** | **0.047** | **0.006** |
| **original_glrlm_RunLengthNonUniformity** | **0.011** | **0.073** | **0.007** |
| **original_glszm_HighGrayLevelZoneEmphasis** | **0.236** | **0.398** | **0.008** |
| **original_ngtdm_Busyness** | **0.020** | **0.127** | **0.009** |
| **original_glszm_GrayLevelNonUniformityNormalized** | **0.079** | **0.189** | **0.011** |
| **original_glcm_Autocorrelation** | **0.388** | **0.903** | **0.014** |
| **original_glcm_JointAverage** | **0.306** | **0.989** | **0.014** |
| **original_glrlm_HighGrayLevelRunEmphasis** | **0.357** | **0.917** | **0.014** |
| **original_glszm_SizeZoneNonUniformity** | **0.086** | **0.100** | **0.018** |
| **original_gldm_DependenceNonUniformity** | **0.037** | **0.108** | **0.019** |
| **original_shape_MajorAxisLength** | **0.087** | **0.243** | **0.022** |
| **original_shape_LeastAxisLength** | **0.135** | **0.229** | **0.023** |
| **original_glrlm_ShortRunHighGrayLevelEmphasis** | **0.495** | **0.771** | **0.024** |
| **original_shape_Maximum2DDiameterColumn** | **0.031** | **0.061** | **0.025** |
| **original_ngtdm_Strength** | **0.039** | **0.083** | **0.027** |
| **original_shape_Maximum3DDiameter** | **0.084** | **0.229** | **0.028** |
| **original_glrlm_GrayLevelNonUniformity** | **0.053** | **0.185** | **0.031** |
| **original_glszm_LowGrayLevelZoneEmphasis** | **0.056** | **0.338** | **0.031** |
| **original_gldm_GrayLevelNonUniformity** | **0.095** | **0.241** | **0.031** |
| **original_glszm_LargeAreaLowGrayLevelEmphasis** | **0.135** | **0.433** | **0.033** |
| **original_firstorder_Range** | **0.011** | **0.023** | **0.034** |
| **original_shape_SurfaceArea** | **0.067** | **0.158** | **0.037** |
| **original_firstorder_Maximum** | **0.009** | **0.026** | **0.037** |
| **original_glszm_SmallAreaEmphasis** | **0.328** | **0.953** | **0.038** |
| **age** | **0.161** | **0.255** | **0.038** |
| **original_gldm_HighGrayLevelEmphasis** | **0.369** | **0.930** | **0.039** |
| **original_shape_Sphericity** | **0.446** | **0.621** | **0.042** |
| **original_ngtdm_Coarseness** | **0.052** | **0.136** | **0.042** |
| **original_glszm_ZoneEntropy** | **0.027** | **0.019** | **0.046** |
| original_shape_Maximum2DDiameterSlice | 0.084 | 0.246 | 0.051 |
| original_glcm_SumEntropy | 0.110 | 0.109 | 0.058 |
| original_firstorder_MeanAbsoluteDeviation | 0.006 | 0.015 | 0.063 |
| original_firstorder_Variance | 0.007 | 0.016 | 0.063 |
| original_glcm_DifferenceVariance | 0.495 | 0.771 | 0.063 |
| original_gldm_LargeDependenceLowGrayLevelEmphasis | 0.236 | 0.819 | 0.065 |
| original_shape_MeshVolume | 0.061 | 0.162 | 0.066 |
| original_shape_VoxelVolume | 0.061 | 0.162 | 0.066 |
| original_firstorder_90Percentile | 0.007 | 0.017 | 0.075 |

| | | | |
|---|---|---|---|
| original_firstorder_InterquartileRange | 0.002 | 0.009 | 0.077 |
| original_firstorder_RobustMeanAbsoluteDeviation | 0.004 | 0.012 | 0.077 |
| original_glrlm_LongRunHighGrayLevelEmphasis | 0.255 | 0.070 | 0.080 |
| original_firstorder_RootMeanSquared | 0.010 | 0.020 | 0.090 |
| original_glcm_DifferenceEntropy | 0.759 | 0.828 | 0.106 |
| original_firstorder_Uniformity | 0.154 | 0.047 | 0.118 |
| original_firstorder_Energy | 0.007 | 0.012 | 0.120 |
| original_firstorder_TotalEnergy | 0.007 | 0.012 | 0.120 |
| original_glcm_Correlation | 0.311 | 0.547 | 0.122 |
| original_glszm_LargeAreaEmphasis | 0.113 | 0.346 | 0.122 |
| original_glszm_ZoneVariance | 0.121 | 0.343 | 0.122 |
| original_firstorder_Kurtosis | 0.057 | 0.023 | 0.127 |
| original_firstorder_Mean | 0.014 | 0.022 | 0.128 |
| original_shape_Maximum2DDiameterRow | 0.140 | 0.335 | 0.132 |
| original_glszm_LargeAreaHighGrayLevelEmphasis | 0.138 | 0.210 | 0.140 |
| original_gldm_DependenceEntropy | 0.210 | 0.430 | 0.148 |
| original_glcm_Idmn | 0.785 | 0.953 | 0.189 |
| original_shape_MinorAxisLength | 0.026 | 0.081 | 0.195 |
| original_gldm_SmallDependenceHighGrayLevelEmphasis | 0.510 | 0.673 | 0.195 |
| original_glcm_Contrast | 0.716 | 0.881 | 0.213 |
| original_glcm_MaximumProbability | 0.236 | 0.097 | 0.229 |
| original_glcm_ClusterTendency | 0.171 | 0.481 | 0.256 |
| original_glrlm_RunEntropy | 0.345 | 0.236 | 0.265 |
| original_firstorder_10Percentile | 0.105 | 0.063 | 0.272 |
| original_glcm_Imc2 | 0.540 | 0.841 | 0.274 |
| original_firstorder_Entropy | 0.194 | 0.063 | 0.295 |
| original_glrlm_ShortRunLowGrayLevelEmphasis | 0.295 | 0.682 | 0.301 |
| original_glszm_ZonePercentage | 0.227 | 0.450 | 0.310 |
| original_ngtdm_Contrast | 0.829 | 0.989 | 0.314 |
| original_gldm_SmallDependenceEmphasis | 0.919 | 0.719 | 0.347 |
| original_glcm_JointEntropy | 0.265 | 0.071 | 0.348 |
| original_glrlm_RunVariance | 0.179 | 0.355 | 0.348 |
| original_glrlm_LongRunEmphasis | 0.236 | 0.457 | 0.352 |
| original_glcm_JointEnergy | 0.28 | 0.073 | 0.375 |
| original_glrlm_ShortRunEmphasis | 0.642 | 0.890 | 0.382 |
| original_glcm_DifferenceAverage | 0.964 | 0.894 | 0.391 |
| original_glrlm_LowGrayLevelRunEmphasis | 0.265 | 0.784 | 0.391 |
| original_firstorder_Median | 0.021 | 0.021 | 0.400 |
| original_glcm_ClusterShade | 0.525 | 0.745 | 0.424 |
| original_gldm_LowGrayLevelEmphasis | 0.250 | 0.736 | 0.448 |
| original_ngtdm_Complexity | 0.594 | 0.749 | 0.452 |
| original_glcm_Imc1 | 0.650 | 0.678 | 0.472 |
| original_glcm_Idn | 0.919 | 0.823 | 0.473 |
| original_glrlm_GrayLevelNonUniformityNormalized | 0.141 | 0.049 | 0.475 |
| original_glrlm_RunLengthNonUniformityNormalized | 0.609 | 0.948 | 0.489 |
| original_shape_Flatness | 0.751 | 0.819 | 0.494 |
| original_gldm_SmallDependenceLowGrayLevelEmphasis | 0.510 | 0.574 | 0.513 |
| original_shape_Elongation | 0.400 | 0.417 | 0.542 |

| | | | |
|---|---|---|---|
| original_gldm_LargeDependenceHighGrayLevelEmphasis | 0.642 | 0.379 | 0.549 |
| original_glrlm_RunPercentage | 0.937 | 0.841 | 0.572 |
| original_glcm_Id | 0.625 | 0.741 | 0.637 |
| original_glszm_GrayLevelVariance | 0.357 | 0.762 | 0.661 |
| original_firstorder_Skewness | 0.838 | 0.536 | 0.721 |
| original_glcm_ClusterProminence | 0.547 | 0.971 | 0.740 |
| original_glszm_SmallAreaHighGrayLevelEmphasis | 0.525 | 0.814 | 0.740 |
| original_glcm_SumSquares | 0.357 | 0.485 | 0.747 |
| original_gldm_LargeDependenceEmphasis | 1.000 | 0.823 | 0.774 |
| original_glcm_Idm | 0.777 | 0.784 | 0.795 |
| original_firstorder_Minimum | 0.407 | 0.452 | 0.858 |
| original_gldm_DependenceNonUniformityNormalized | 0.982 | 0.665 | 0.910 |
| original_gldm_GrayLevelVariance | 0.301 | 0.346 | 0.926 |
| original_glrlm_GrayLevelVariance | 0.400 | 0.510 | 0.983 |
| original_gldm_DependenceVariance | 0.601 | 0.780 | 0.983 |

**Supplementary Table 4.** Log-rank test for continuous MFS.

| | | N = 115 | p-value (continuous MFS) |
|---|---|---|---|
| Sex | MALE | 83 | 0.714 |
| | FEMALE | 32 | |
| HP | SQUAMOUS | 77 | 0.835 |
| | NONSQUAMOUS | 38 | |
| Location | LEFT | 65 | 0.536 |
| | RIGHT | 50 | |
| T | T12 | 41 | 0.917 |
| | T34 | 74 | |
| N | N01 | 25 | 0.656 |
| | N23 | 90 | |
| Zubrod score | ZERO | 34 | 0.818 |
| | NONZERO | 81 | |

### Supplementary Figures

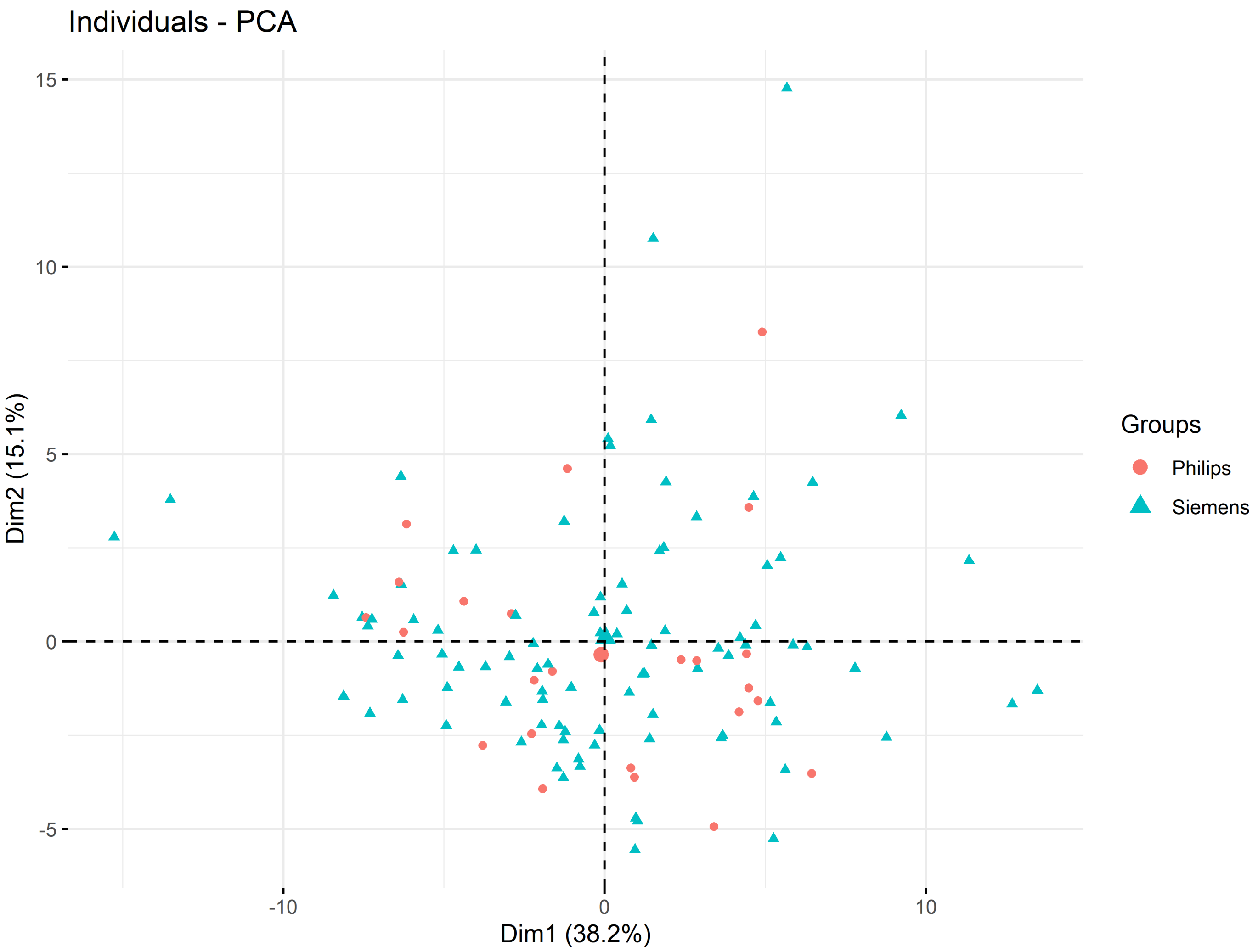


**SF 1.** Principal component analysis of the radiomic features (after correlation filtering). Colors correspond to the PET/CT scanner. There is no visible grouping of samples according to the scanner.

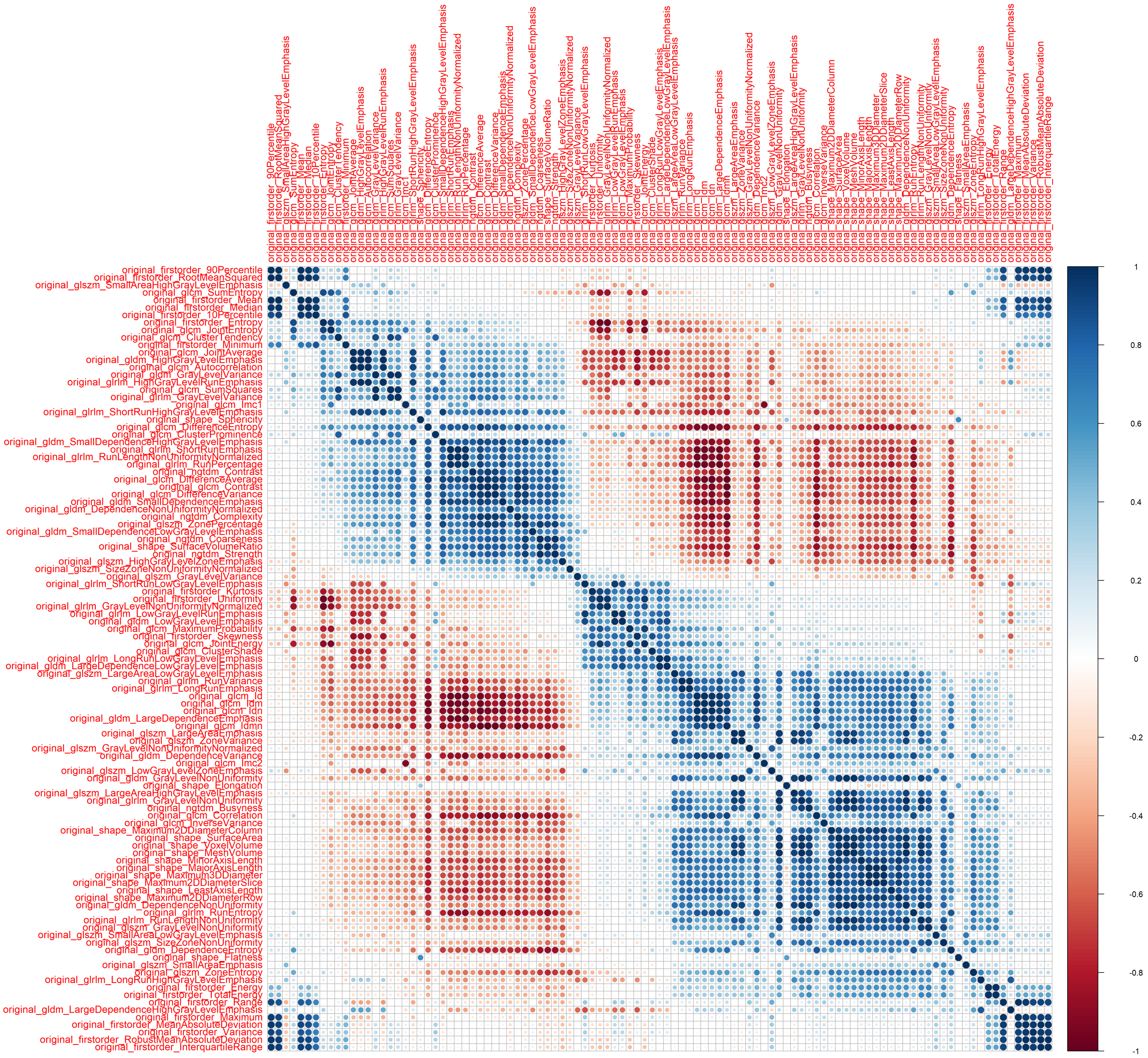


**SF 2.** Correlation between radiomic features.

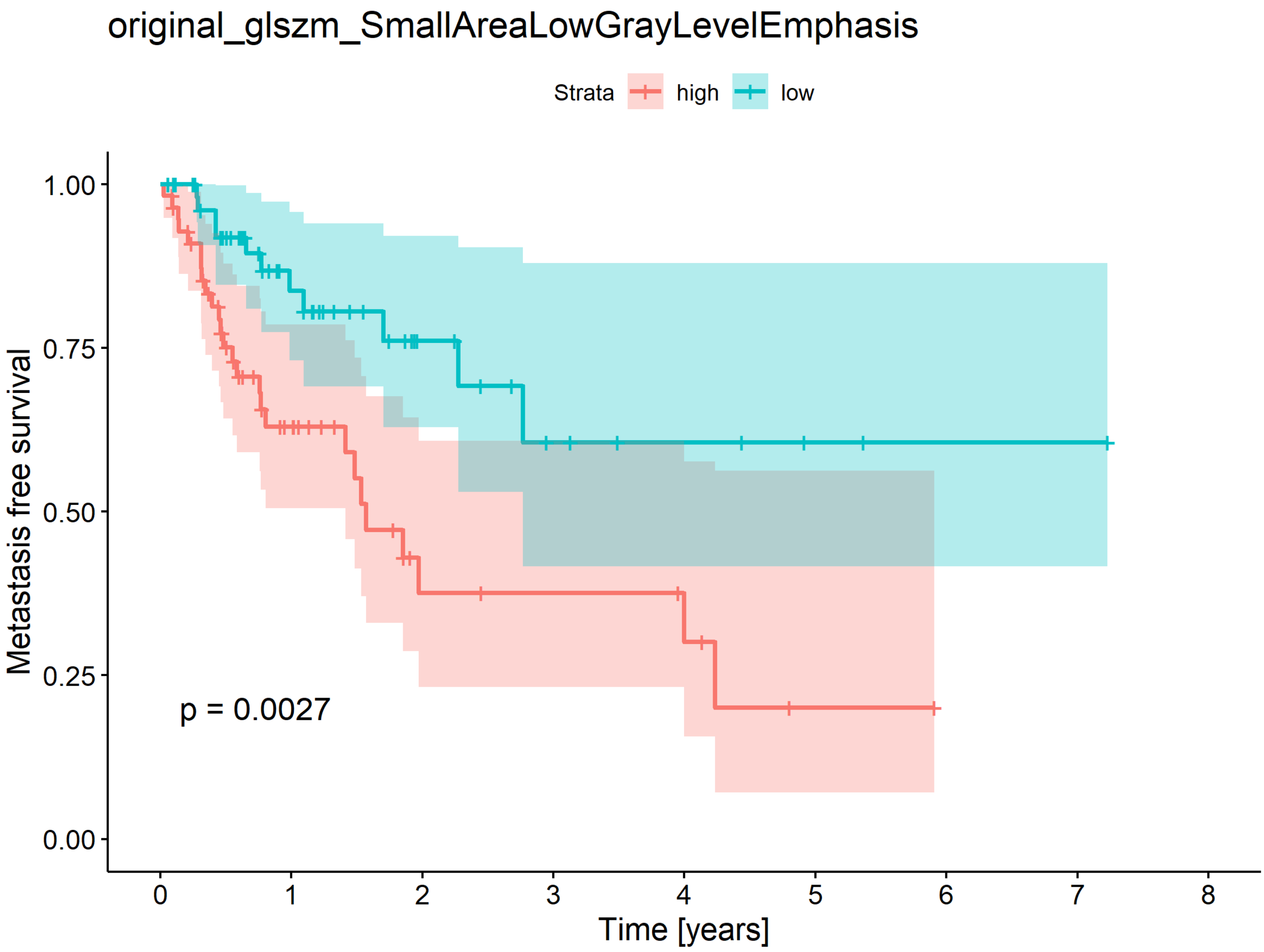


**SF 3.** Kaplan-Meier plot for metastatic-free survival with high/low SmallAreaLowGrayLevelEmphasis value

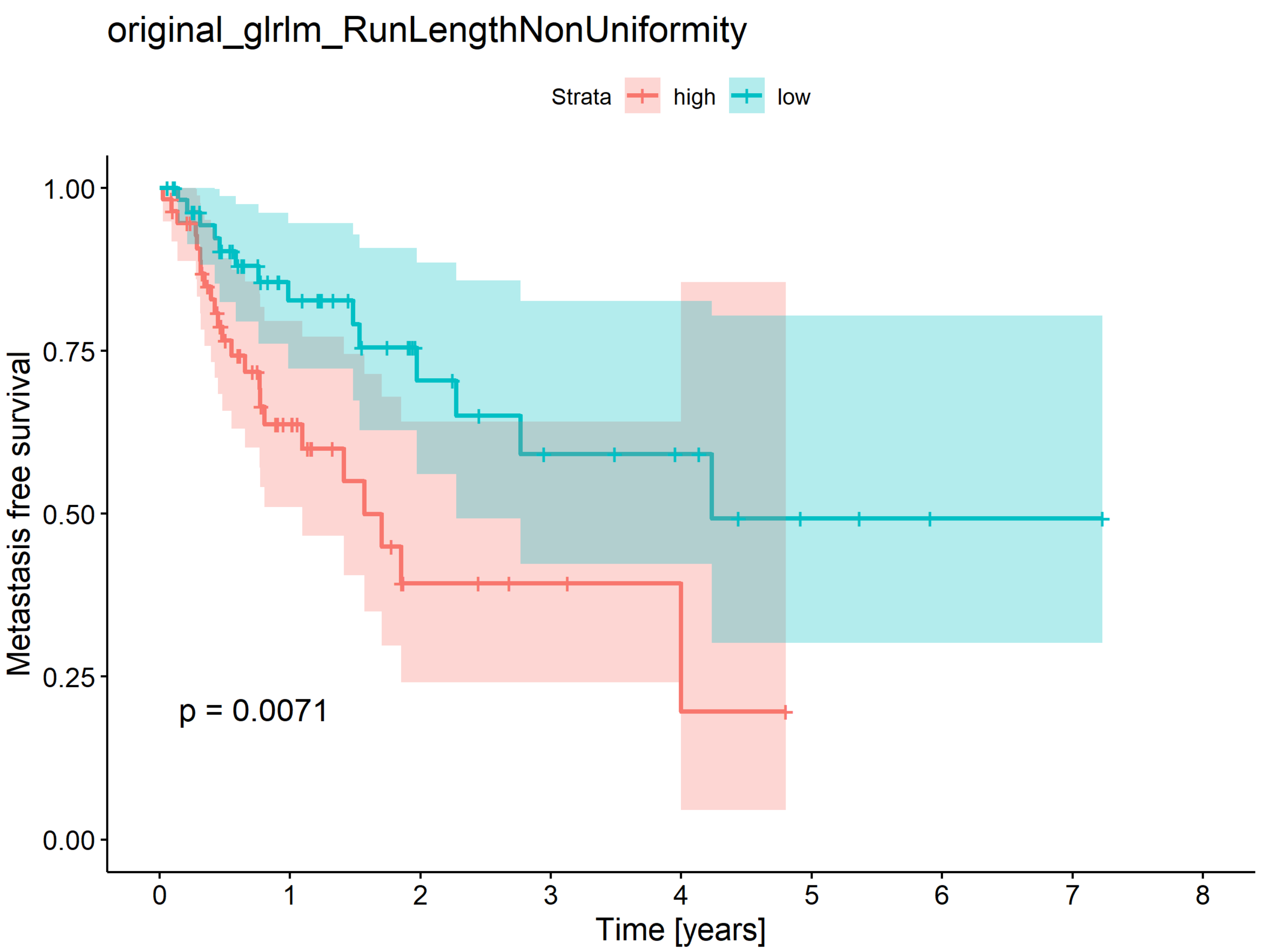


**SF 4.** Kaplan-Meier plot for metastatic-free survival with high/low RunLengthNonUniformity value
